# Mapping routine measles vaccination and factors related to uncompleted vaccination in Japan: An ecological study

**DOI:** 10.1101/2021.10.23.21265425

**Authors:** Yuki Matsumoto

## Abstract

**Objective:** Few studies in Japan have considered regional differences in immunisation coverage and have geographically examined its distribution. We aimed to identify the municipal-level distribution of immunisation coverage and explore regional characteristics associated with its regional differences through ecological studies.

**Design:** An ecological study.

**Setting:** Japan municipalities.

**Participants:** All 1737 municipalities.

**Outcome measure:** The mean measles-rubella (MR) vaccination rate distribution from 2013 to 2018 was drawn using GIS. We tested the null hypothesis of non-random distribution using spatial autocorrelation analysis (Global Moran’s I). We presented clusters of municipalities with significantly higher and lower vaccination rates by hot spot analysis (Getis-Ord Gi*). Furthermore, we estimated among municipality characteristics, regional ones associated with vaccination rates.

**Results:** According to Global Moran’s I, the MR vaccine coverage distribution was clustered significantly (z = 11.44, p <0.00). Based on the hot spot analysis, the western Japan region tended to have more cold spots with low inoculation rates, small numbers of births and areas with considerable annual variations in vaccination rates. In multiple regression analysis using regional characteristics as independent variables, the 1-year and 6-month check-up rates (Model 2: 95% confidence interval [CI], 0.21-0.36), number of births (0.96-1.54), and standard deviation of the vaccination rate in a target year per municipality (0.17-0.24), were positively associated with the vaccination rate. Contrariwise, average mothers’ age (−1.15 to -0.34), percentage of single-parent households (−0.48 to -0.17), percentage of households with multiple children (−0.12 to -0.04), and moving-in rates (−0.26 to -0.05) were negatively associated with vaccination rate.

**Conclusion:** To eliminate regional differences in immunisation coverage, it is practical to identify areas with low immunisation coverage for a proactive corrective approach. The regional characteristics identified in this study, such as population structure, administrative characteristics, and trends in socioeconomic status in municipalities will make it possible to identify target areas.

**Strengths and limitations of this study:**

- The mean measles-rubella (MR) vaccination rate distribution from 2013 to 2018 was drawn using GIS.
- We tested the null hypothesis of non-random distribution using spatial autocorrelation analysis (Global Moran’s I).
- We presented clusters of municipalities with significantly higher and lower vaccination rates by hot spot analysis (Getis-Ord Gi*).
- Furthermore, we estimated among municipality characteristics, regional ones associated with vaccination rates

## INTRODUCTION

Measles is a highly contagious disease, and in 2018, more than 140,000 people died from measles, most of whom were children under the age of five.[1] However, measles is an infectious disease that can be prevented by vaccination, and the number of deaths has been decreasing worldwide with the introduction of vaccination.[2] In Japan, measles vaccines became available and vaccination has been implemented since the 1960s, and the current measles-rubella (MR) combination vaccine was introduced in 2006.[3] The number of measles infections in Japan has also been decreasing due to the widespread use of vaccines, and the World Health Organisation (WHO) recognised Japan’s measles elimination status in 2015.[4] On the other hand, dozens of imported cases have been reported in Japan from 2015 to 2020.[4,5] The spread of infectious diseases has become easier in today’s globalised society, and it is necessary to identify barriers and encourage more vaccinations in Japan.

It is well-known that multiscale regional disparities exist in immunisation coverage. The WHO has set a goal of achieving a 90% or higher coverage rate for the first dose of measles vaccine in each country and an 80% or higher coverage rate at the subnational level.[1] First, in terms of global disparities, countries and regions in the Global South (e.g., India, Lebanon, Nigeria, Venezuela) tend to have lower coverage rates.[6] In comparison, countries and regions in the Northern Hemisphere have higher coverage rates. On a more micro-scale, regional differences have been shown in studies examining the distribution of vaccination coverage among regions in Asia, such as Bangladesh,[7] Mongolia,[8] and Indonesia.[9] Studies examining geographic differences in immunisation coverage include those that determined association of immunisation coverage with either individual-level characteristics (maternal age and education),[10,11] or regional-level factors (urban versus suburban areas,[12,13] access to good health care providers,[14] and the state of social infrastructure in areas of residence).[8] These regional characteristics may influence vaccine hesitancy, which refers to “delay in acceptance or refusal of vaccination despite availability of vaccination services”[15], and the World Health Organisation (WHO) identified the phenomenon as a threat to global health in 2019.[16] The decision-making of vaccination is an individual’s behaviour, but it is also related to broader social contexts such as the historical, political, and socio-cultural contexts.[17]

Few existing studies in Japan have examined the distribution of vaccination coverage across multiple regions. In Japan, there has been an increasing number of studies examining the influence of individual-level variables on vaccination behaviour at the regional level (e.g., level of mother’s knowledge and attitudes toward vaccination,[18,19] and concerns about adverse reactions).[20,21] Also, studies have confirmed association of household-level characteristics of sample populations with either vaccination behaviour or household attributes (e.g., income[22] and the number of children in the household).[23,24] These studies tended to focus only on a single region or community. In recent years, some studies have been conducted to examine factors at the regional level, including the use of multilevel models, showing associations of vaccination behaviour with social mobility,[25] nationality,[25] and the number of paediatricians in the region.[26] Although the Ministry of Health, Labour, and Welfare (MHLW) has compiled municipal vaccination rates for each prefecture,[27] these have not been projected on a map for geographical analysis. The Strategic Advisory Group of Experts on Immunisation established by the WHO has developed a strategy, set to be achieved by 2030, advocating national and subnational monitoring.[28] Understanding vaccination coverage at the municipal level is an important issue in terms of its contribution to public and global health.

Therefore, based on the hypothesis that regional differences in immunisation coverage exist in Japan as in other countries, this study aimed to identify the distribution of immunisation coverage at the municipal level and explore regional characteristics associated with regional immunisation coverage differences in this ecological study. Combined with individual-level findings of previous studies, these study findings may enable tailor-made vaccination policies at the administrative unit levels, and more efficient implementation of vaccination programmes.

## METHODS

### Data collection

#### Vaccination rate

Vaccination rates for the MR vaccine (Phase 1), a routine immunisation, were reported by the MLWH. The municipality vaccination rates, from 2013 to 2018, included in this study were published by the MLWH.[27] The choice of these target years of the data collection was based on the year the law on immunisation was revised (2013), and the year of the latest data (2018), which were available as of May 2021. However, in this study, the analysis was conducted using the average vaccination coverage rates in the target years. In addition, the goal of this study was to project regional differences rather than track the yearly variations in vaccination coverage; this is because panel data are unavailable for most of the covariates. Finally, for municipalities that underwent administrative unit changes or mergers within the years of interest, we processed the data to correspond to the 2018 administrative classification (two regional mergers occurred in 2014). One town, each in 2016 and 2018, was upgraded to a city because of population growth.

#### Independent variables

Variables suggested to be significantly related to immunization coverage in previous studies were mainly retrieved from government statistical databases. The definitions of these variables are listed in Table 1. For vaccination rate, when data were released over time, we used the average value within the target years for the analysis; however, for data that were released just once from 2013 to 2018, we used the data for that single year (Table 1). Municipalities with missing explanatory variable values were excluded from the analysis. The following variables were included in the analysis for each municipality: average mothers’ age at child’s birth (between 2014 and 2018),[18,21,22,24,29] percentage of households with single parents,[25] (2015) per capita income(log)[22-24] (between 2013 and 2018), infant health check-up rate[31] (2013–2018), percentage of households with working mothers[22,24,25] (2015), moving-in rate[25] (2013–2018), percentage of births with non-Japanese nationalities[25,30,31] (2013–2018), percentage of households with more than one child[23,24,29,32] (2015), number of births(log) (2013–2018), standard deviation of vaccination coverage, the number of medical institutions offering MR vaccination, and the number of paediatricians[22,33] (2014, 2016, 2018). These 12 variables were entered into the regression model as covariates. The models that included the number of medical institutions for MR vaccination and the number of paediatricians were designated as Models 1 and 2, respectively. These two variables indicate accessibility and could cause multicollinearity. Except for medical institutions offering MR vaccination, all the variables were retrieved from the governmental statistical databases. The sources were as follows:

**Table 1.** Variables selected as candidates in the regression models

| Variables | Definition |
| --- | --- |
| Vaccination coverage | Percentage of the population taking MR vaccination (first dose) |
| Standard deviation of vaccination coverage | Standard deviation of vaccination coverage between 2013 and 2018 |
| Average mothers' age at child's birth | Average age of the mothers when giving birth |
| Per capita income(log) | Log-transformed income per number of registered residents |
| Percentage of households with single parent | Percentage of the households with single-parent (0-5 years old) |
| Percentage of households with working mother | Percentage of the households with working mothers (0-2 years old) |
| Percentage of households with more than one child | Percentage of the households with more than one child (0-2 years old). |
| Infant health check-up rate | Percentage of the 18-month infant population taking infant health check-up |
| Percentage of births with non-Japanese nationalities | Percentage of new-born children whose nationality are non-Japanese |
| Moving-in rate | Number of people (0-4 years old) moving into the municipalities divided by the total population (0-4 years old) of the municipalities |
| Number of births(log) | Log-transformed yearly number of new-born children |
| Number of paediatricians | Number of paediatricians in the municipalities |
| Number of hospitals offering MR vaccination | Number of hospitals offering MR vaccination |

MR, measles-rubella

MHLW: infant health check-up[33], the number of paediatricians[34], average mothers’ age,[34] and number of births,[35]

Ministry of Internal Affairs and Communications: per capita income,[36] working mothers,[37] nationalities,[38] moving-in rate,[38] single-parent rate,[37] and more than one child rate.[37]

### Quantitative analysis

#### Spatial analysis

Immunisation rates were projected at the municipality level. We then tested for spatial autocorrelation by Global Moran’s I, using the k-nearest neighbour method (k=8) to examine whether the geographic distributions were random or had clusters. Then, if the distribution was non-random, we attempted to identify geographic areas with low vaccination rates using hot spot analysis (Getis-Ord Gi*), by the k-nearest neighbour method (k=8). In hot spot analysis, for statistically significant positive Z-scores, the higher the Z-score, the stronger the clustering of higher values (hot spots). For statistically significant negative Z-scores, the smaller the Z-score, the stronger the clustering of lower values (cold spots). The standard deviation of the vaccination rate per municipality for the entire target year and the number of births that approximated the distribution of the target population were also projected on the map. Furthermore, geocoding was performed using the addresses of medical institutions corresponding to MR vaccines, and the number of point features per municipality was recorded as the number of medical institutions. Ver. 2.5 of ArcGIS Pro (Esri Inc., California, United States) was used for these analyses. For the municipal boundary data, the National City Boundary Data, version 8.2.1, published by Esri Japan (Tokyo, Japan) (https://www.esrij.com/products/japan-shp/), retrieved on 6 December 2020, were used following some processing.

### Statistical analysis

Multiple regression analysis using the forced imputation method was conducted for municipalities. Municipalities with missing variable values were excluded from the analysis. In addition, to avoid multicollinearity, variables with a variance inflation factor (VIF) >10 were analysed by removing one of them from the model. For this multivariate analysis, we used R Studio version 1.3.959 (R version 4.0.00). The level of significance was set a priori at .05.

### Patient and public involvement statement

Patients and public were not involved in the design study.

## RESULTS

### Geographical trends

In the analysis of immunisation coverage, all 1737 municipalities were included in the study. The national average vaccination coverage was 95.19% (Figure.1). Spatial autocorrelation analysis using Global Moran’s I statistics at a significance level of 95% and 9999 permutations showed a significantly higher vaccination coverage distribution in the eastern part of Japan than in the western part. (z-score = 11.44, Moran’s index = 0.12). Hot spot analysis using k-nearest neighbour values (k=8) also showed a tendency for more cold spots distribution in western Japan. The distribution in Figure 2 is similar to the distribution of hot spots (Figure 3), which are areas with a high standard deviation of vaccination coverage in the target year. The distribution in Figure 3 is also similar to the distribution in areas with low births (Figure 4). Areas with a small geographic distribution of medical facilities offering MR vaccine tended to have particularly high and particularly low immunisation rates (Figure 5).

**Figure 1.**
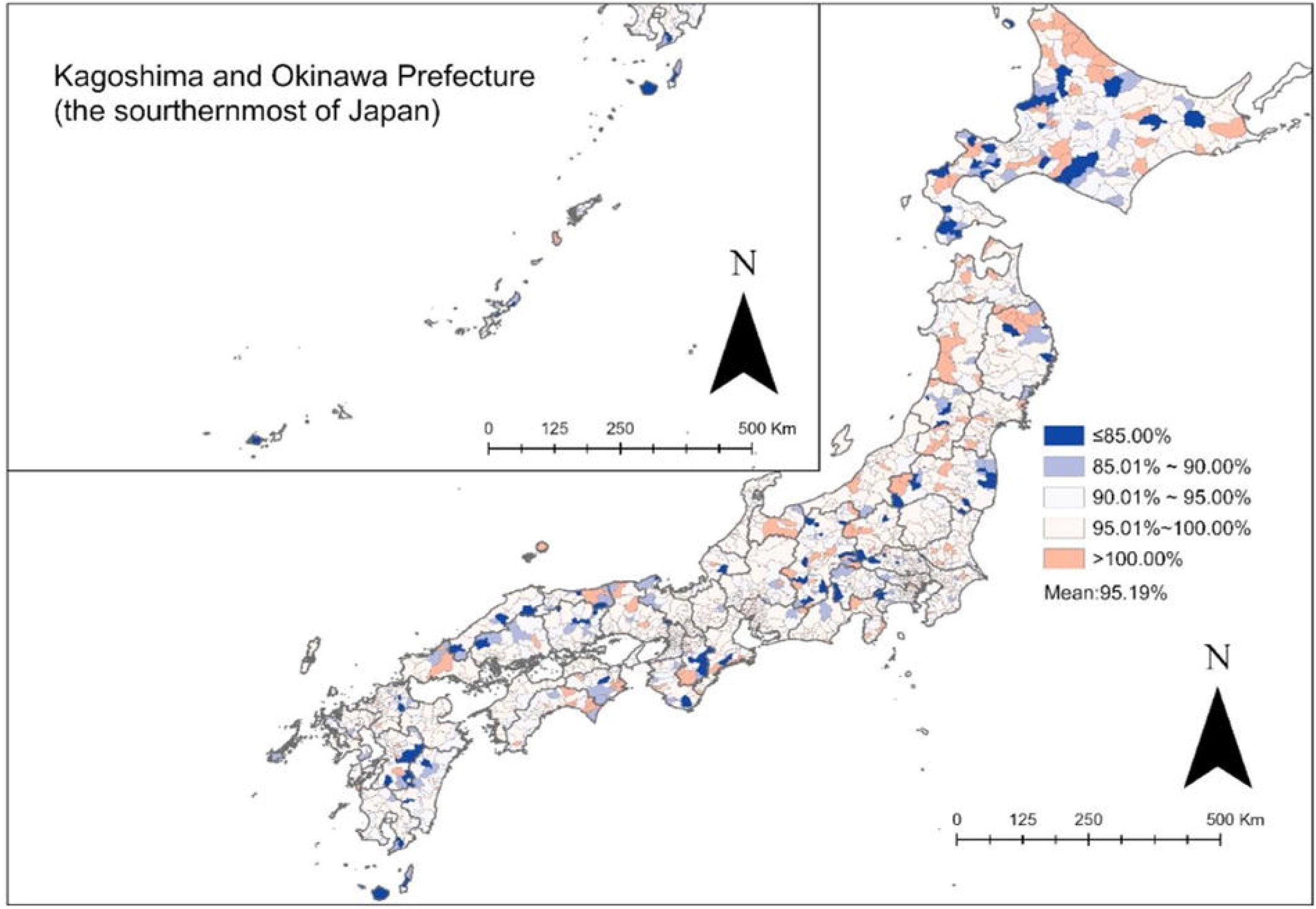
Spatial distribution of MR vaccination coverage by municipalities. MR, measles-rubella

**Figure 2.**
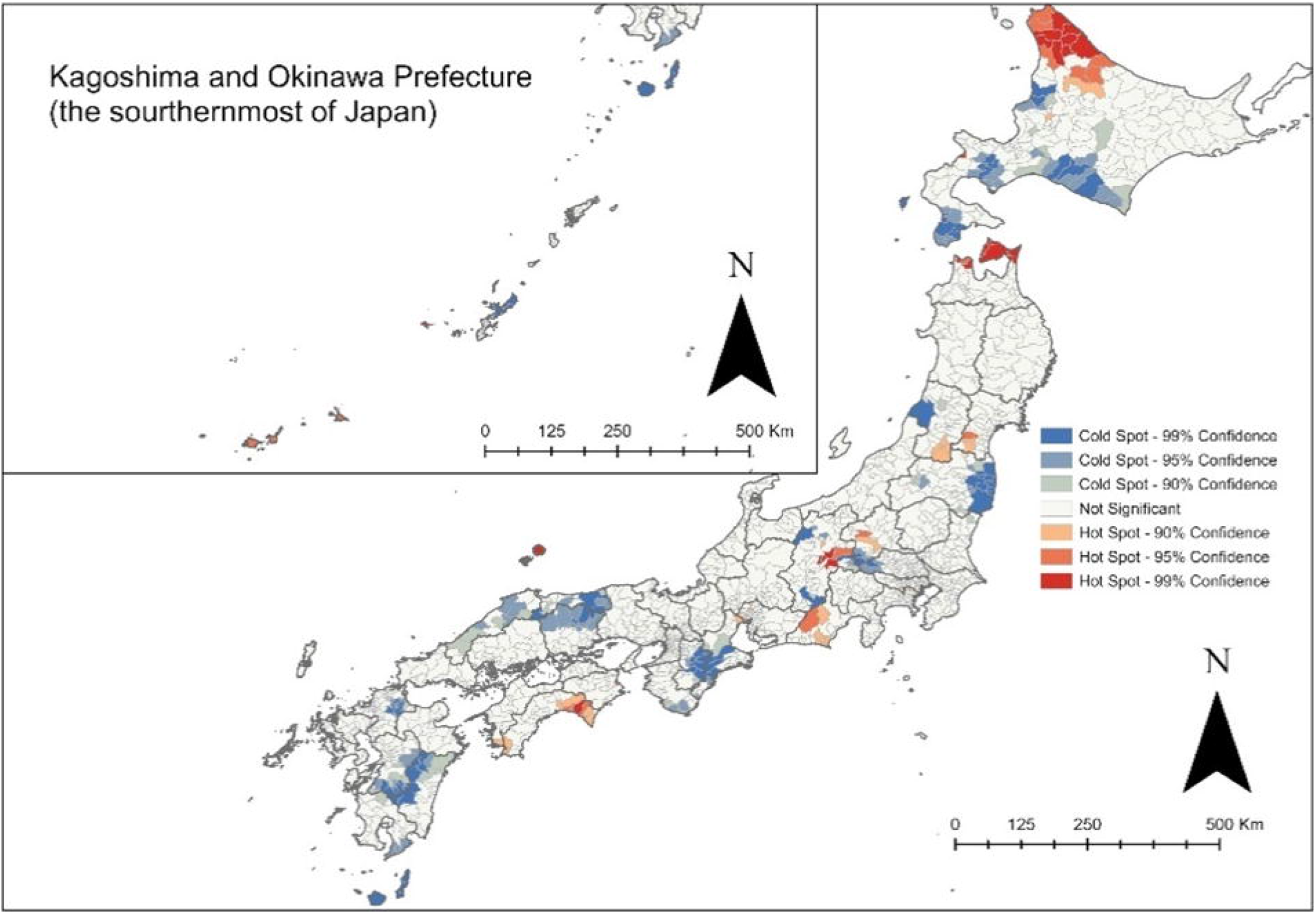
Mapping hot and cold spot analysis of MR vaccination coverage by k-nearest neighbour algorithm (k =8) MR, measles-rubella

**Figure 3.**
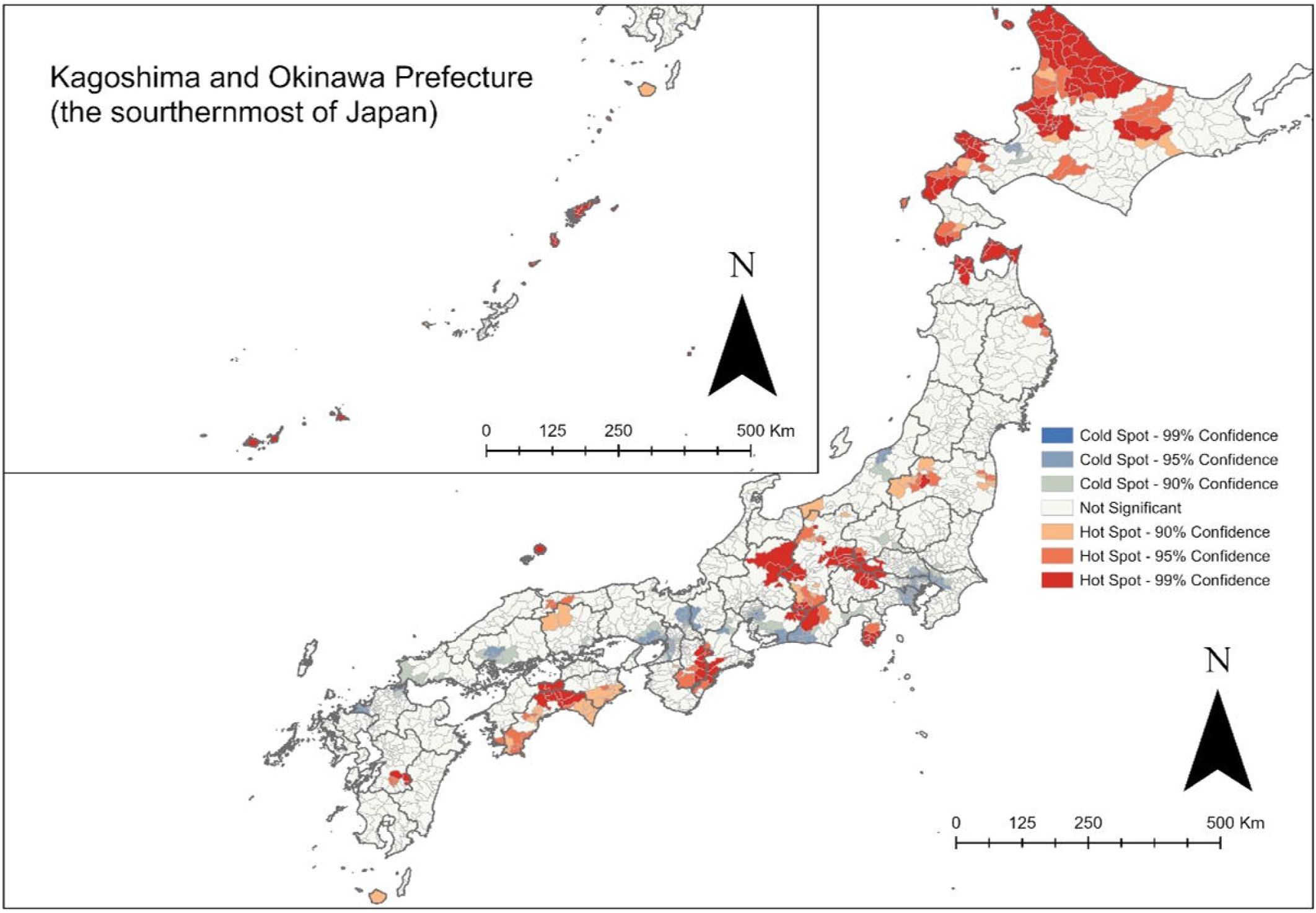
Mapping hot and cold spot analysis of MR vaccination coverage standard deviation by k-nearest neighbour algorithm (k =8). MR, measles-rubella

**Figure 4.**
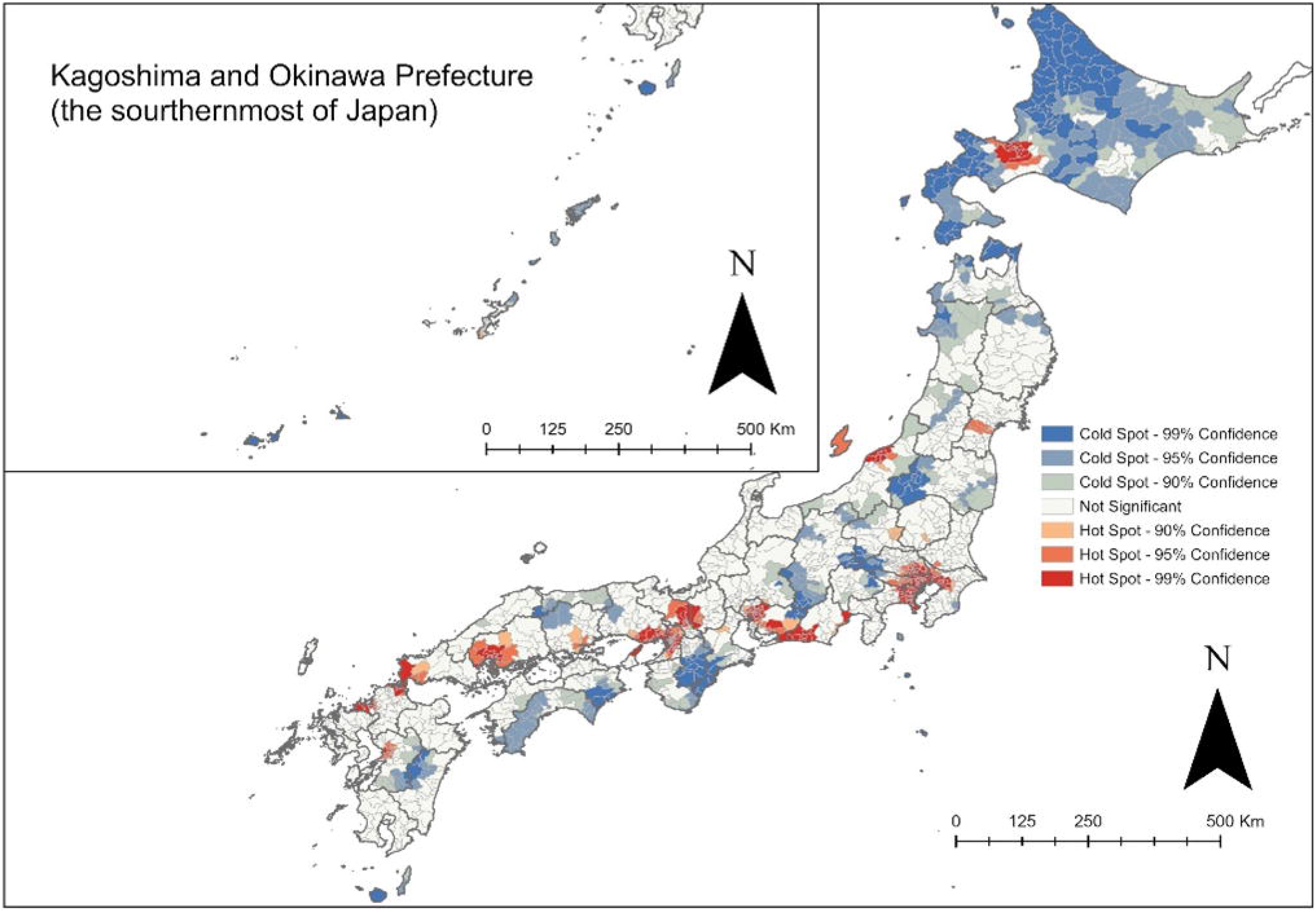
Mapping hot and cold spot analysis of number of new-born children by k-nearest neighbour algorithm (k = 8).

**Figure 5.**
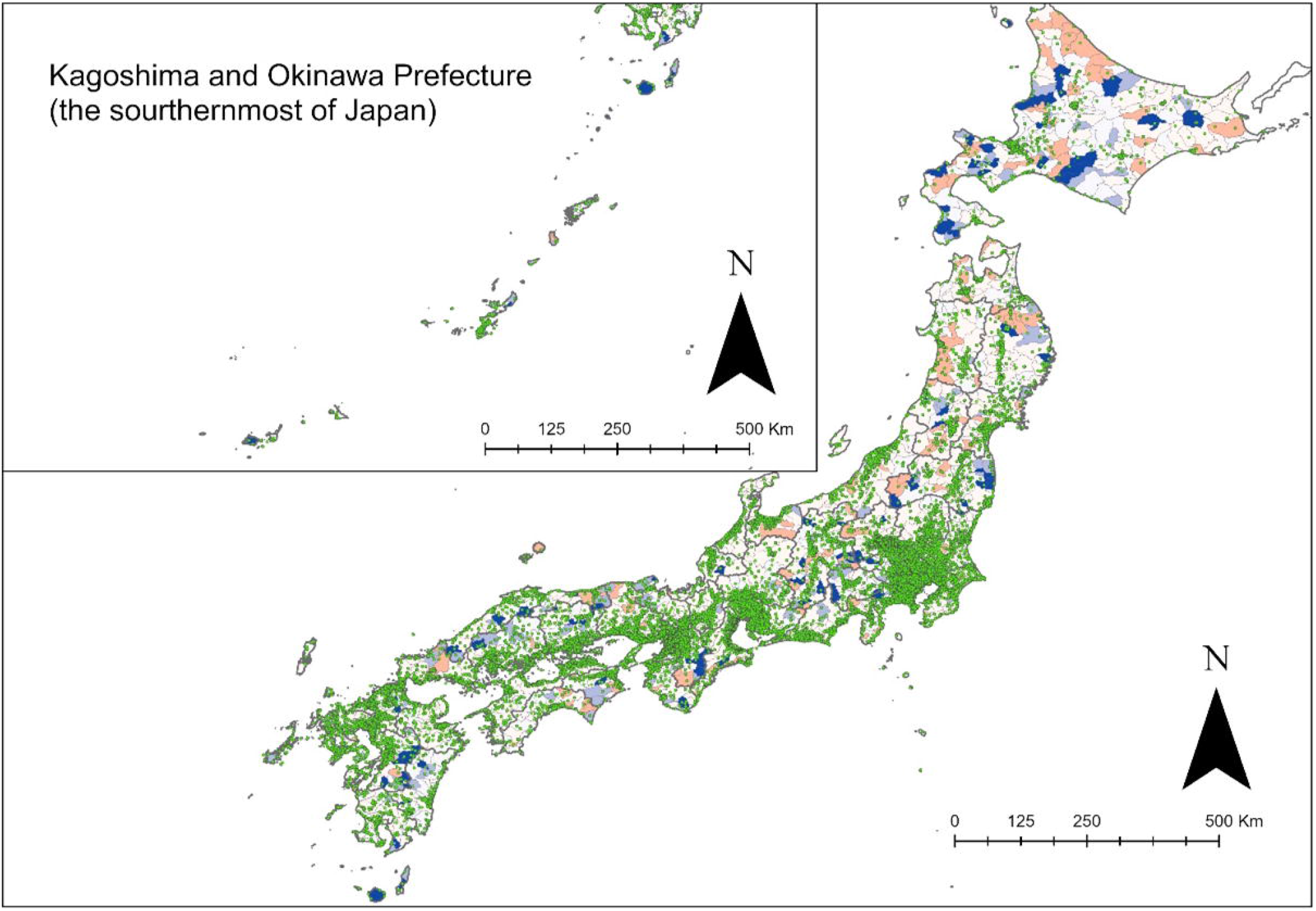
Spatial distribution of hospital offering MR vaccination (green dots) on the map of MR vaccination coverage by municipalities. MR, measles-rubella

### Statistical trends

After excluding regions with missing values in the dataset of regional characteristics, the total number of municipalities included in the analysis was 1731. The intraclass correlation coefficient for immunisation coverage when 1731 municipalities were nested in 47 prefectures was <0.20 (0.03, 95% confidence interval [CI], 0.01-0.06). Therefore, we did not consider the two administrative levels (the prefecture and municipality). The means and standard deviations of the variables used in the statistical analyses are presented in Table 2. The national average vaccination coverage was 95.25%, and its mean value was then subjected to a general multiple regression analysis using the forced imputation method. To avoid multicollinearity, the multiple regression model included the number of paediatricians per municipality as the accessibility variable in Model 1. Multiple regression analysis included the number of medical institutions that can provide MR vaccine per municipality as the accessibility variable in Model 2. The results of the analyses are presented in Table 3. The variables that showed significant associations in Models 1 and 2, were the same. In both models, the VIF function for determining multicollinearity among the independent variables was <10. In Models 1 and 2, mothers’ age (95% CI, -1.16-0.34 vs -1.15-0.34), percentage of single-parent households (−0.48 -0.17 vs -0.48-0.17), percentage of households with multiple children (−0.12-0.04 vs -0.12-0.04), 1-year-and 6-month check-up rate (0.21-0.37 vs 0.21 0.36), excess moving-in rate (−0.26 to 0.05 vs -0.26 -0.05), number of births (0.93-1.51 vs 0.96-1.54), and standard deviation of vaccination coverage between 2013 and 2018 (0.17-0.24 vs 0.17-0.24), respectively, were significantly associated. In both Models 1 and 2, accessibility variables (number of healthcare providers offering MR vaccine and number of paediatricians) were not significantly associated.

**Table 2.**
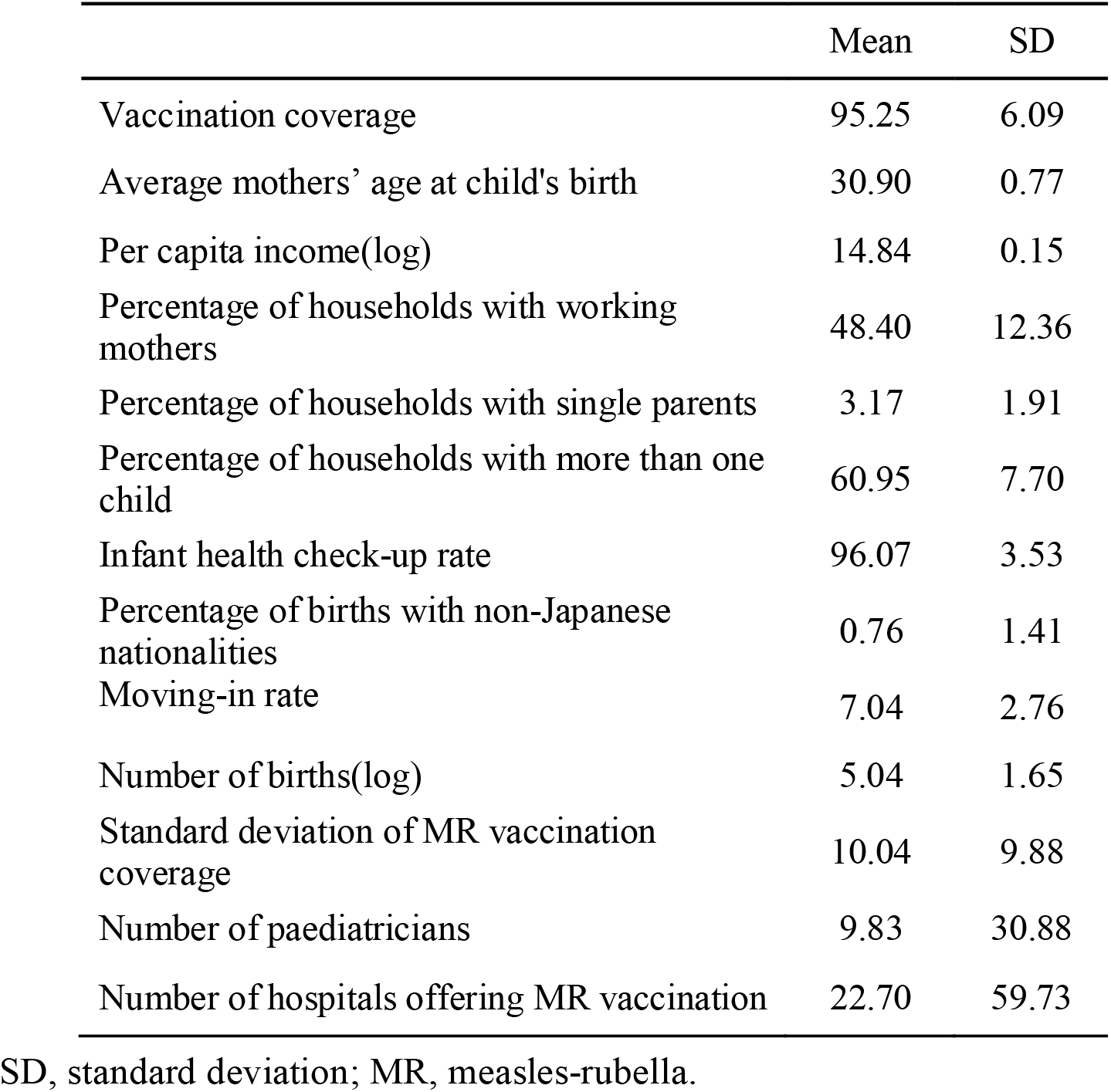
Descriptive statistics using municipality as a unit of analysis

|  | Mean | SD |
| --- | --- | --- |
| Vaccination coverage | 95.25 | 6.09 |
| Average mothers' age at child's birth | 30.90 | 0.77 |
| Per capita income(log) | 14.84 | 0.15 |
| Percentage of households with working mothers | 48.40 | 12.36 |
| Percentage of households with single parents | 3.17 | 1.91 |
| Percentage of households with more than one child | 60.95 | 7.70 |
| Infant health check-up rate | 96.07 | 3.53 |
| Percentage of births with non-Japanese nationalities | 0.76 | 1.41 |
| Moving-in rate | 7.04 | 2.76 |
| Number of births(log) | 5.04 | 1.65 |
| Standard deviation of MR vaccination coverage | 10.04 | 9.88 |
| Number of paediatricians | 9.83 | 30.88 |
| Number of hospitals offering MR vaccination | 22.70 | 59.73 |

SD, standard deviation; MR, measles-rubella.

**Table 3.** Results from the multivariate regression model with municipality (n= 1731)

|  | Model 1 |  |  |  |  | Model 2 |  |  |  |  |
| --- | --- | --- | --- | --- | --- | --- | --- | --- | --- | --- |
|  | Estimate | Std. Error | 95% CI |  | P value | Estimate | Std. Error | 95% CI |  | P value |
| Average mothers' age at child's birth | -0.75 | 0.21 | -1.16 | -0.34 | 0.00*** | -0.75 | 0.21 | -1.15 | -0.34 | 0.00*** |
| Per capita income(log) | 2.19 | 1.39 | -0.53 | 4.91 | 0.11 | 2.18 | 1.38 | -0.54 | 4.89 | 0.12 |
| Percentage of households with working mothers | 0.00 | 0.01 | -0.02 | 0.04 | 0.45 | 0.01 | 0.01 | -0.02 | 0.04 | 0.46 |
| Percentage of households with single parents | -0.33 | 0.08 | -0.48 | -0.17 | 0.00*** | -0.33 | 0.08 | -0.48 | -0.17 | 0.00*** |
| Percentage of households with more than one child | -0.08 | 0.02 | -0.12 | -0.04 | 0.00*** | -0.08 | 0.02 | -0.12 | -0.04 | 0.00*** |
| Infant health check-up rate | 0.29 | 0.04 | 0.21 | 0.37 | 0.00*** | 0.29 | 0.04 | 0.21 | 0.36 | 0.00*** |
| Percentage of births with non-Japanese nationalities | 0.05 | 0.10 | -0.15 | 0.26 | 0.61 | 0.06 | 0.10 | -0.15 | 0.26 | 0.59 |
| Moving-in rate | -0.15 | 0.05 | -0.26 | -0.05 | 0.01** | -0.15 | 0.05 | -0.26 | -0.05 | 0.01** |
| Number of births(log) | 1.22 | 0.15 | 0.93 | 1.51 | 0.00*** | 1.22 | 0.15 | 0.96 | 1.54 | 0.00*** |
| Standard deviation of MR vaccination coverage | 0.21 | 0.19 | 0.17 | 0.24 | 0.00*** | 0.21 | 0.18 | 0.17 | 0.24 | 0.00*** |
| Number of paediatricians | -0.01 | 0.00 | -0.02 | 0.00 | 0.12 |  |  |  |  |  |
| Number of hospitals offering MR vaccination |  |  |  |  |  | -0.01 | 0.00 | -0.01 | 0.00 | 0.06 |
| Intercept | 56.73 | 20.37 | 16.78 | 96.69 | 0.00** | 56.74 | 20.33 | 16.86 | 96.62 | 0.01** |
| Adjusted R squared | 0.16 |  |  |  |  | 0.16 |  |  |  |  |

\* p<0.05, \*\* p<0.01, \*\*\*p<0.001. Std. Error, standard error; CI, confidence interval; MR, measles-rubella

## DISCUSSION

The national average MR vaccination rate in Japan was 95.18% (Table 1). This rate was 10% higher than the global first dose average of 86%.[1] Similar to the results of previous studies conducted in other countries,[6-13] the results suggest regional clusters of low and high vaccination coverage areas in Japan (Figure 1). The distributions of hot spots (with high vaccination rates) and cold spots (with low vaccination rates) were similar to the annual variations in the number of births and vaccination rates (standard deviation of vaccination rates in the target year) and the distribution of areas with few medical institutions that provide MR vaccines (Figures 3-5). To examine whether these distributional trends were significant even when controlling for other regional characteristics, we used a general multiple regression model with 12 variables (Models 1 and 2). The results showed that there was no significant association of accessibility to paediatricians and medical institutions providing MR vaccines with vaccination coverage. Among the regional characteristics, the 1-year and 6-month check-up rates, number of births, and standard deviation of the vaccination rate in a target year per municipality, were positively associated with the vaccination rate. On the other hand, the average age of mothers, percentage of single-parent households, percentage of households with multiple children, and moving-in rates were negatively associated with vaccination rate.

The geographic distribution of vaccination rates in western and eastern Japan may reflect the tendency of many sparsely populated towns and villages in the mountainous regions of western Japan. However, evidence suggests that there is no clear distance-attenuation trend in vaccination rate differences between rural and urban areas,[39] and this study did not show a clear geographical trend in the distribution of vaccination rates. In addition to psychological factors, political, cultural, and social factors are also associated with vaccination hesitancy,[40] and it is possible that some socio-cultural and political characteristics were common to these spots. The lower the number of births in an area, the lower the number of people eligible for vaccination. The fewer the number of births, the greater the variation in vaccination coverage. If a town has 100 people eligible for vaccination and only one person does not get vaccinated, the town’s vaccination rate will be 99%. In a community with 5 people eligible for vaccination, even if only 1 person fails to be vaccinated, the vaccination rate for that area would be 80%.

In statistical analysis, four variables were negatively associated with vaccination coverage. Previous studies in other countries using individual-level data have suggested that children from single-parent families[41,42] and, a finding that is consistent with these reports. Single parents tend to be disadvantaged in terms of socioeconomic status compared to families to which both parents belong. Several studies have reported low immunisation coverage after the second child.[<u>42]</u> Different acquisition of information on child rearing depend on the child’s birth order, and complacency may also have an impact.[43] Regarding the age of the mothers, the results are inconsistent with those of several countries.[9,44] Japan has a low birth rate,[45] and the age at first marriage and first childbirth is tending towards increase.[46,47] In this context, areas with a high number of births to those younger than the average age(30.9 years old) can be the focus of active governmental implementing measures on child rearing (e.g., subsidies and grants for childcare, expansion of nurseries, and provision of child rearing courses). Thereby, the government may actively recommend vaccinations for infants. The fact that the transfer rate was negatively associated with vaccination is inconsistent with the results of a previous study in Japan.[25] In addition to the complexity of the procedure, it may be difficult for the local government to make timeous recommendations to new residents regarding whether they wished to be vaccinated in a different area than the one in which they were notified of the vaccination. It is also possible that the vaccination process affected the method used to calculate the number of people eligible for vaccination, as infants are socially transferred to new administrative areas.

There are three variables that have shown positive associations in statistical analysis. These results are consistent with those of previous studies that have found a positive association between health check-ups and timely immunisation.[31 In Japan, infant health check-ups at one year and six months of age include items on completed or incomplete immunisation, and health check-ups can be used to encourage those who are yet unimmunised. This may suggest a relationship with vaccination rates at the regional level. The following reason may explain why the number of births suggested a positive association with the magnitude of annual variations in vaccination rates. the areas with the lowest number of births are concentrated in non-urban mountainous areas, where the shortage of medical personnel and the aging of the population are issues.[48,49] The aging of the population as a whole and the associated decline in the working-age population may have an impact on health care administration. Furthermore, as mentioned earlier, variations in vaccination rates tend to be greater in areas with fewer births. A small number of non-vaccinated people in sparsely populated areas may have contributed to a large drop in vaccination coverage.

Five variables showed no association. Unlike previous studies, the results of this study did not show an association between income and vaccination coverage.[6] Income inequality at the municipal level may not be as strongly associated with decisions to vaccinate as socioeconomic status at the individual and household levels. This study did not find a significant association with working mothers. This may be due to a mixture of factors, such as single-mother households and dual-earner households, which have different socioeconomic periodical conditions. For example, it has been suggested that parental leave has a positive impact on vaccination.[20] The percentage of foreign births was also not significantly related to the number of children. The presence or absence of an ethnic community of foreigners residing in the area and the diversity of their cultural characteristics, as well as differences in the extent to which information and services are provided in foreign languages by the government in different areas, may have had a bidirectional effect on the vaccination rate. The variables related to accessibility did not show a significant association with immunisation coverage in this study. It is possible that challenges may arise in the immunisation supply system in the future. The lack of a significant association with paediatricians may be due in part to the fact that non-paediatricians can provide immunisations in Japan.[32] In the case of influenza vaccination in the elderly, evidence showed that vaccination rates increased with proximity to the facilities where vaccination was available.[50] For infant vaccination, both mothers and children are younger than the elderly, and convenience in terms of distance may thus be less of a barrier to vaccination.

### Limitations

First, the data on vaccination rates were incomplete. In addition to the yearly rates, average vaccination rates above 100% were found in this analysis. One of the reasons for this is that the vaccination rate is calculated by dividing the number of vaccinated persons at a certain point in time by the number of vaccinations conducted in the target year, and the rate does not consider the results of population movement. Previous studies have revealed difficulties in accurate ascertainment of vaccination coverage.[43] There is a need to obtain accurate data on the vaccination coverage of individual residents.

Second, there is a possibility of ecological fallacy. Trends at the regional level do not necessarily correspond to individual trends. The challenge is that residents in regions with unique variable values related to the vaccination rates do not necessarily have unique characteristics. Furthermore, panel data could not be used in this study. For the yearly vaccination rates, we used the average value and attempted to reduce the yearly variations, but some of the variables were only available for a single year, and the yearly variations may have significantly affected the distribution of the variables. Moreover, the results of the analysis in this study were not able to prove causality, although we reported the correlations.

## CONCLUSION

This study examined the geographic distribution of vaccination rates in Japan, assuming that differences in vaccination rates exist between regions, as in other countries. As a result, although there appeared to be no clear clusters in vaccination rates at first glance, statistical analysis (Global Moran’s I) suggested that clusters existed, with many low areas distributed in the west. Related to the difference in vaccination rates was the demographic structure of the municipality, the percentage of families with inferior socioeconomic status, and the possible influence of government childcare policies. There are relatively few studies on vaccine hesitancy in Asia and even fewer in Japan. Factors of vaccine hesitancy depend on the type of vaccination and the local context. As vaccine hesitancy factors depend on the type of vaccination and the local context, more studies focusing on Japan and Asia are needed. More efficient immunisation policies will be possible if the results of this study and previous studies are examined on a more refined regional scale and through panel data analysis covering a more extended period. To achieve this, it will be necessary for the government to further strengthen its information management system for individual-level vaccination information, and that the method for calculating vaccination coverage be re-examined.

## Data Availability

All data produced in the present study are available upon reasonable request to the authors

## Acknowledgments

This study was based on the author’s master’s thesis. We would like to thank Department of Geography, Graduate School of Letters, Kyoto University.

## Author Contributions

YM was involved in study design, data interpretation, data analysis and wrote the manuscript.

## Patient consent for Publication

Not required.

## Provenance and peer review

Not commissioned; externally peer-reviewed.

## Research Ethics approval

No formal ethical approval was required.

## Data sharing statement

Data are from e-stat (Japanese government) and medley, Esri Japan. The dataset can also be requested by emailing the corresponding author.

## Open Access

This is an open access article distributed in accordance with the Creative Commons

## Attribution Non-Commercial (CC BY-NC 4.0) license

which permits others to distribute, remix, adapt, build upon this work non-commercially, and license their derivative works on different terms, provided the original work is properly cited and the use is non-commercial. See http://creativecommons.org/licences/by-nc/4.0/

© Article author(s) (or their employer(s) unless otherwise stated in the text of the article) 2021. All rights reserved. No commercial use is permitted, unless otherwise expressly granted.

